# Unveiling the oncogenic significance of thymidylate synthase in human cancers

**DOI:** 10.1101/2024.01.05.24300913

**Authors:** Yibo Geng, Luyang Xie, Yang Wang, Yan Wang

**Affiliations:** Department of Neurosurgery, Beijing Chaoyang Hospital, Capital Medical University; Department of Neurosurgery, Beijing Tiantan Hospital, Capital Medical University

**Keywords:** thymidylate synthase, tumor, TGCA, prognosis, tumor-infiltrating immune cell

## Abstract

**Background:** Thymidylate synthase (TYMS) constitutes a pivotal and potent target in the context of chemoresistance. However, the oncogenic role of TYMS has received insufficient attention.

**Methods:** Leveraging data from the Cancer Genome Atlas and various public databases, we conducted an extensive investigation into the oncogenic role of TYMS across 33 cancer types.

**Results:** TYMS exhibited pronounced expression across a spectrum of cancers and demonstrated associations with clinical outcomes in diverse cancer patient cohorts. Furthermore, genetic alterations were identified as potential influencers of overall survival in specific tumor types. Notably, the expression of thymidylate synthase correlated with tumor-infiltrating CD4+ cells in select cancers. Additionally, the functional mechanism of TYMS encompassed nucleotidase activity, chromosome segregation, and DNA replication progress.

**Conclusions:** This study furnishes a comprehensive understanding of the oncogenic role played by TYMS in human tumors.

## Introduction

The exploration of novel oncogenes is crucial for achieving a comprehensive understanding of the mechanisms underlying malignant tumors and identifying potential therapeutic targets[1]. Utilizing resources such as The Cancer Genome Atlas and various other publicly available analysis websites, which house valuable genomics and proteomics information[2], facilitates interactive analyses specifically focused on unraveling the oncogenic role of thymidylate synthase (TYMS).

TYMS, a human nucleotide enzyme, plays a pivotal role in endogenous thymidylate synthesis, enabling de novo production of thymidylate. While TYMS has been extensively studied in contexts such as hepatitis, rheumatic diseases, and neural development[3-5], recent investigations have increasingly highlighted its significance in chemoresistance and tumorigenesis[6, 7]. This paper synthesizes findings from laboratory-based experiments involving cell or animal models, elucidating the intricate relationship between TYMS and various malignancies (Figure 1) [8-24].

**Figure 1.**
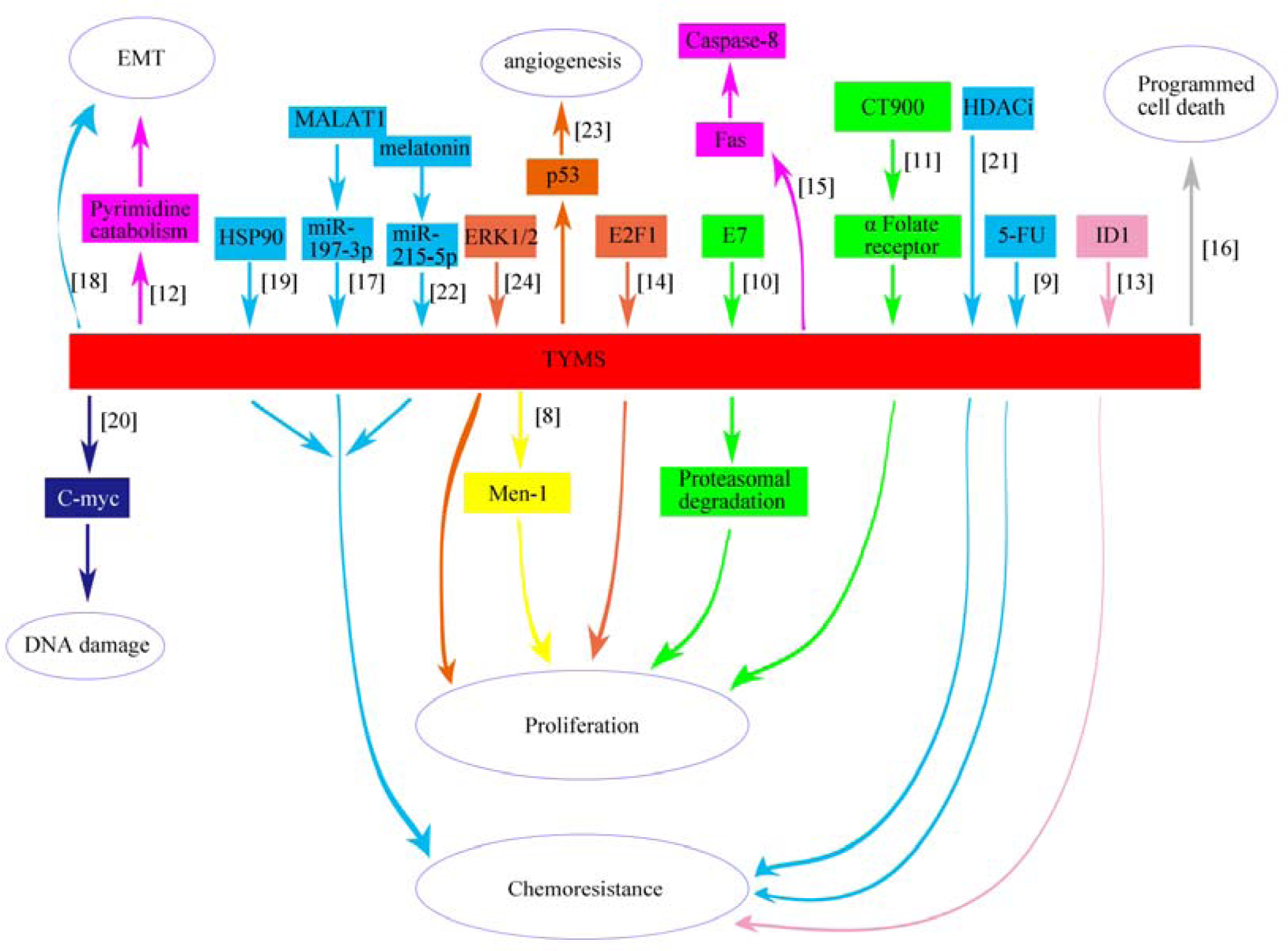
Schematic depicting the relationship between TYMS and various tumors.

In our study, we employed data from The Cancer Genome Atlas and several public databases to analyze oncologic features, encompassing gene expression, survival rates, genetic alterations, phosphorylation sites, immune cell infiltration, and related gene functions. This comprehensive approach aims to provide a detailed understanding of the oncogenic mechanisms governed by TYMS.

## Materials and Methods

### Gene location and protein structure analysis

The genomic location of the TYMS gene was determined based on the UCSC genome browser (GRCh38/hg38) [25]. Utilizing the “HomoloGene” function of the NCBI, conserved functional domain analysis of the TYMS protein across different species was conducted. The three-dimensional structure of TYMS was obtained using the cBioPortal web tool[26].

### Gene expression analysis

#### TIMER2

Differences in TYMS expression were assessed using the “Gene_DE” module of the Tumor Immune Estimation Resource, version 2 (TIMER2) [27]. In cases lacking contrast tissues or a sufficient contrast group, the Gene Expression Profiling Interactive Analysis, version 2 (GEPIA2) web server from Genotype-Tissue Expression (GTEx) was employed[28]. Violin plots depicting TYMS expression across various pathological stages were generated using the “Pathological Stage Plot” module in GEPIA2, utilizing log2 [TPM (Transcripts per million) +1] transformed expression results with a log-scale test.

#### HPA

The Human Protein Atlas (HPA) database provided TYMS expression data in different cells, tissues, and plasma[29]. Plasma sample data were estimated through mass spectrometry-based proteomics, defining “low specificity” as “Normalized expression ≥1 in at least one tissue/cell type, but not elevated in any tissue/cell type.” Immunohistochemistry (IHC) images of TYMS in five pairs of normal and tumor tissues (BRCA, COAD, LIHC, and LUAD) were downloaded from HPA and analyzed.

### Survival analysis

Utilizing the Kaplan-Meier “Survival Map” module in GEPIA2, overall survival (OS) and disease-free survival (DFS) maps for TYMS were obtained. The expression threshold for categorizing high/low expression groups was set at 50%. Kaplan-Meier survival analysis in GEPIA2 generated survival plots using the log-rank test.

### Genetic alteration analysis

Genetic alteration features, alteration frequency, mutation type, and mutated site information for TYMS were collected from the cBioPortal web. Overall survival data for tumors with or without TYMS genetic alterations were collected, and Kaplan-Meier analysis with log-rank P-values was performed.

### Phosphorylation feature

The predicted phosphorylation features of TYMS at sites S6, T53, S114, S124, Y146, S151, Y153, S154, and T167 were obtained from the open-access PhosphoNET database by searching the protein name “TYMS.”

### Immune infiltration cell analysis

The association between TYMS expression and immune infiltrates, specifically CD4+ T-cells, CD8+ T-cells, cancer-associated fibroblasts, and NK cells, was explored using the TIMER2 web tool. Purity-adjusted Spearman’s rank correlation test provided P-values and partial correlation (cor) values.

### TYMS-related gene enrichment

The STRING website was employed to identify TYMS-binding proteins[30], with a low confidence score set at 0.7. Interaction types were based on the maximum number of interactors (≤50), full STRING network, and confidence.

### Data availability

The datasets analyzed in this study are available in the online dataset. Requests for further access to the dataset can be directed to.

## Results

### Gene location and protein structure

In our study, we investigated the oncogenic function of TYMS (NM_001071 or NP_001062.1, Supplementary Figure 1A). The protein structure demonstrated conservation across various species (Supplementary Figure 1B, C).

### Gene expression analysis

The expression levels of TYMS in various normal tissues and blood cells were assessed. The thymus exhibited the highest TYMS expression, followed by bone marrow and tonsil. TYMS displayed a tissue-enhanced (immune tissues) expression pattern, detected in all tissues except skeletal muscle and tongue (Figure 2A). Blood cell analysis showed low specificity, with markedly elevated TYMS expression in regulatory T cells (Figure 2B).

**Figure 2.**
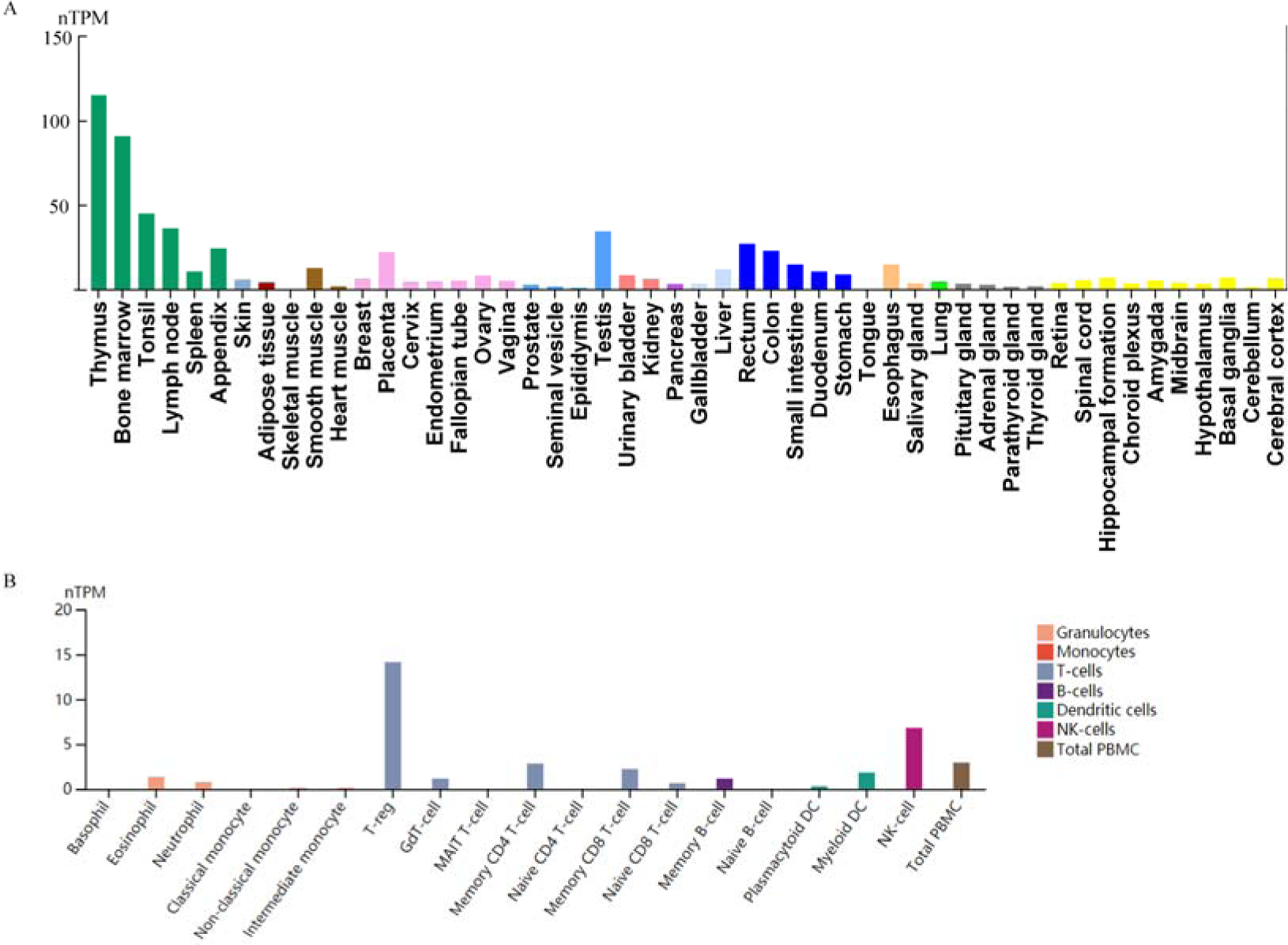
TYMS expression in normal organ. (A) Expression level of TYMS in normal human tissues. Different color indicated different human system. (B) TYMS expression in normal blood immune cells.

TYMS expression in multiple tumors, including BLCA, BRCA, CESC, CHOL, COAD, ESCA, GBM, HNSC, KIRC, KIRP, LIHC, LUAD, LUSC, PCPG,STAD, THCA, and UCEC, was higher than in corresponding normal tissues (Figure 3A, P<0.01). Further analysis using the GTEx dataset revealed significant differences in expression levels between tumor and normal tissue in additional cancers (Figure 3B, P<0.05). However, TYMS expression in KICH, PRAD, TGCT, or THCA was similar to normal tissue (Supplementary Figure 2, P>0.05).

**Figure 3.**
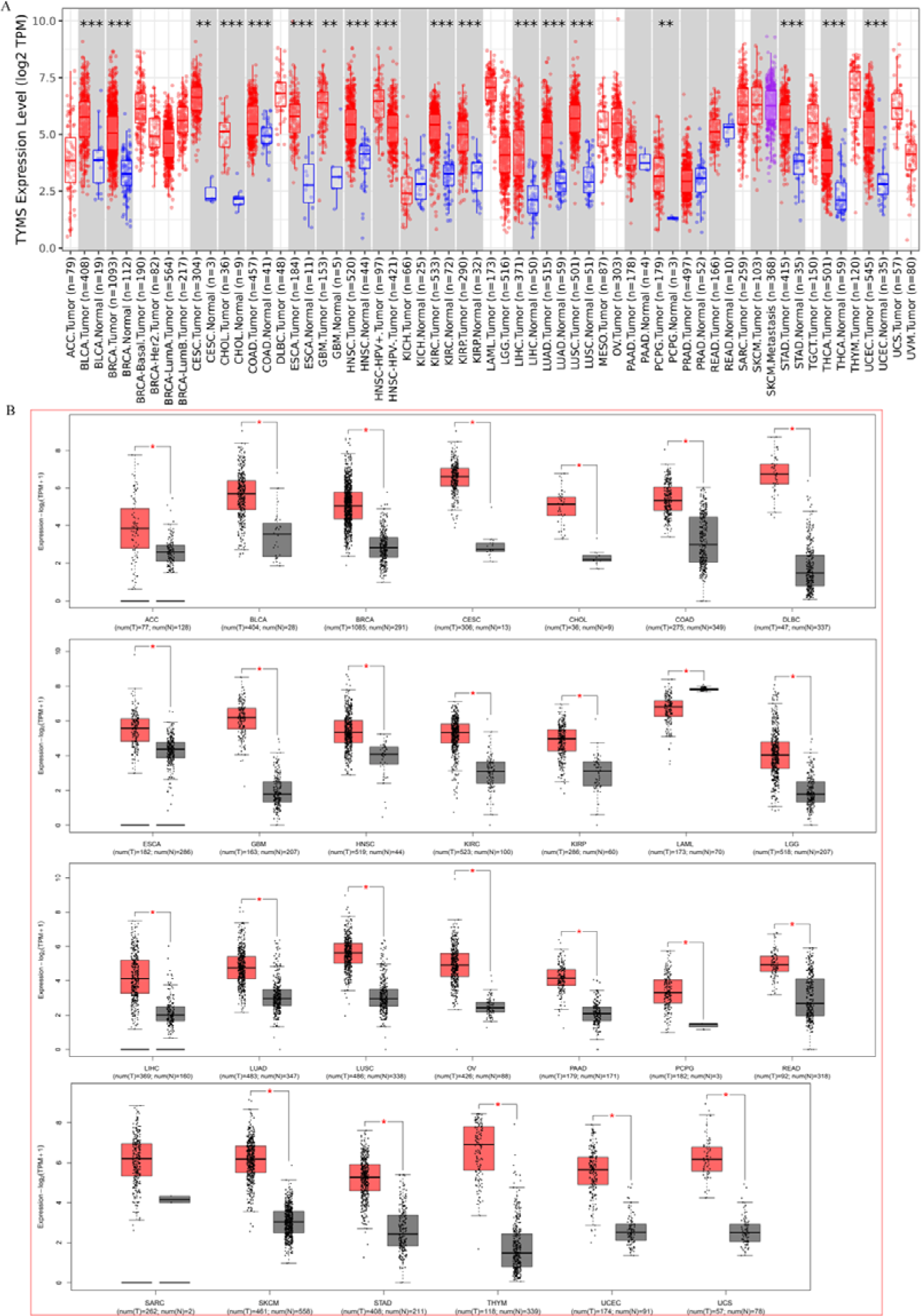
TYMS expression difference across various tumors. (A) TYMS expression between various tumors and comparable normal tissues through TCGA dataset. (B) TYMS expression through TCGA and GTEx dataset. *P < 0.05; **P < 0.01; ***P < 0.001

Positive correlations were found between TYMS expression and pathological stages in ACC, KICH, LIHC, and TGCT. Conversely, COAD, LUSC, and OV showed negative correlations (Figure 4). No correlation was observed in other cancers (Supplementary Figure 3).

**Figure 4.**
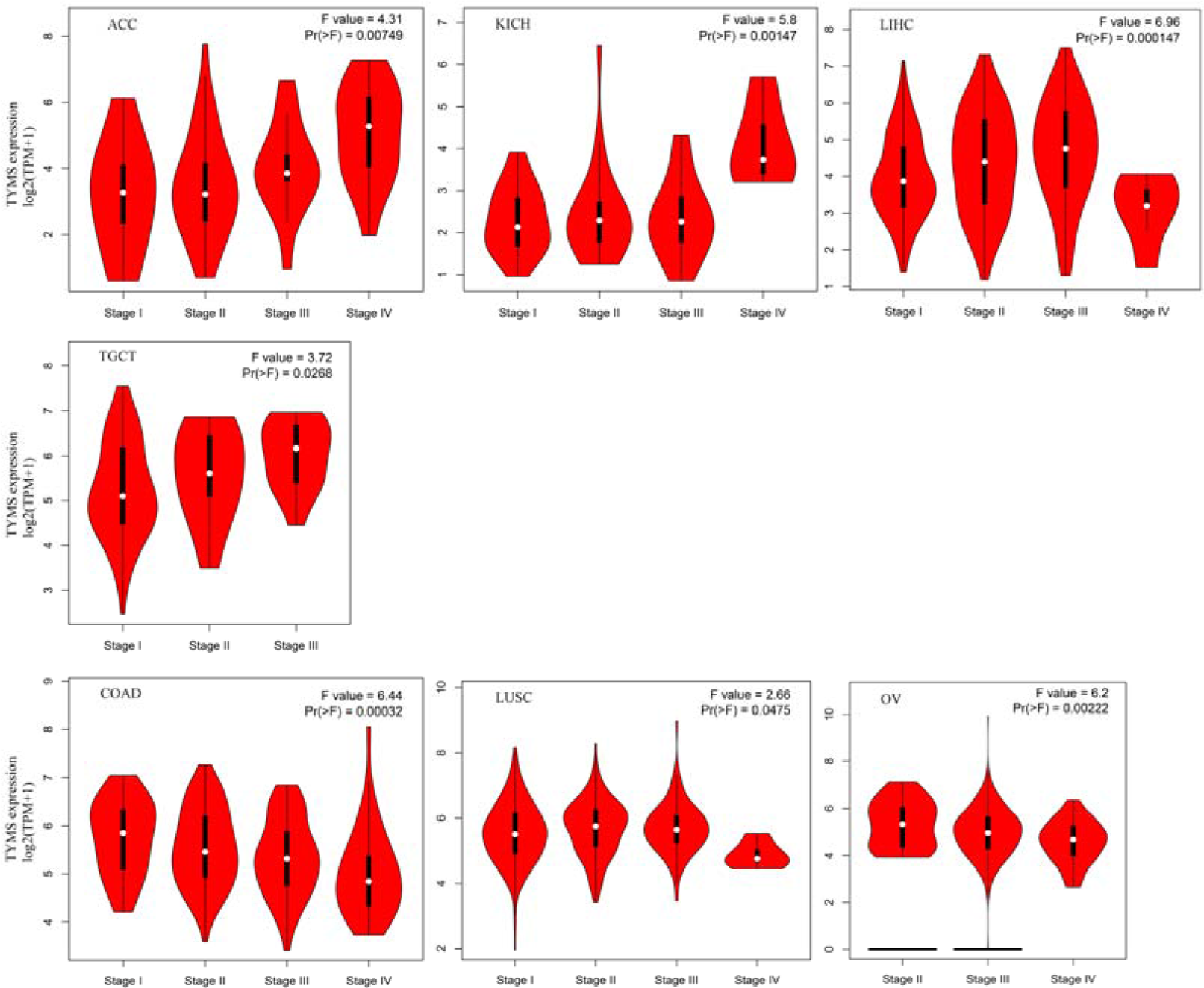
TYMS expression difference across pathological stages in seven tumor types.

### Immunohistochemistry difference

Comparison of TYMS staining between normal and tumor tissues via IHC corroborated TYMS expression patterns in the HPA dataset. Medium or strong TYMS staining was observed in BRCA, COAD, LIHC, and LUAD, while low or negative staining was evident in normal comparable tissues (Figure 5).

**Figure 5.**
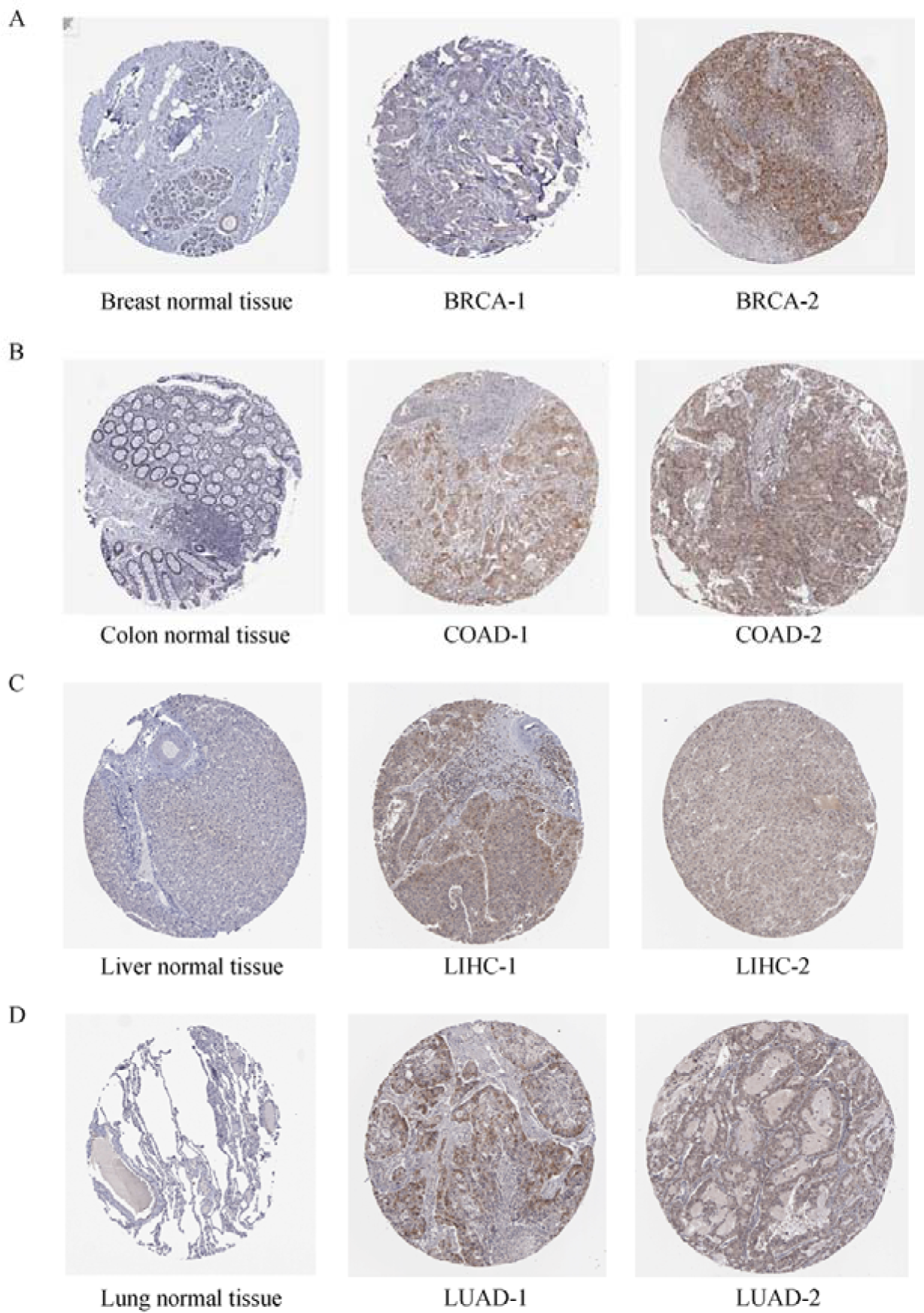
IHC difference between normal and tumor tissues in BRCA (A), COAD (B), LIHC (C) and PRAD (D). (P < 0.05)

### Survival analysis

High TYMS expression correlated with poor overall survival in ACC (P<0.001, HR=3.8), KICH (P<0.05, HR=9.2), LGG (P<0.001, HR=2.1), LIHC (P<0.001, HR=1.8), LUAD (P<0.001, HR=1.7), MESO (P<0.001, HR=3.3) and SARC (P<0.01, HR=1.8) (Figure 6A). Similarly, it was associated with poor DFS in ACC (P<0.001, HR=4.9), KICH (P<0.05, HR=4.9), LGG (P<0.01, HR=1.5), LIHC (P<0.001, HR=1.8), PRAD (P<0.001, HR=2.0) and SARC (P<0.05, HR=1.5) (Figure 6B). While TYMS showed varied associations with outcomes across different cancers, certain cancers, including ACC, KICH, LGG, LIHC and SARC exhibited consistent tendencies in both OS and DFS (Figure 6).

**Figure 6.**
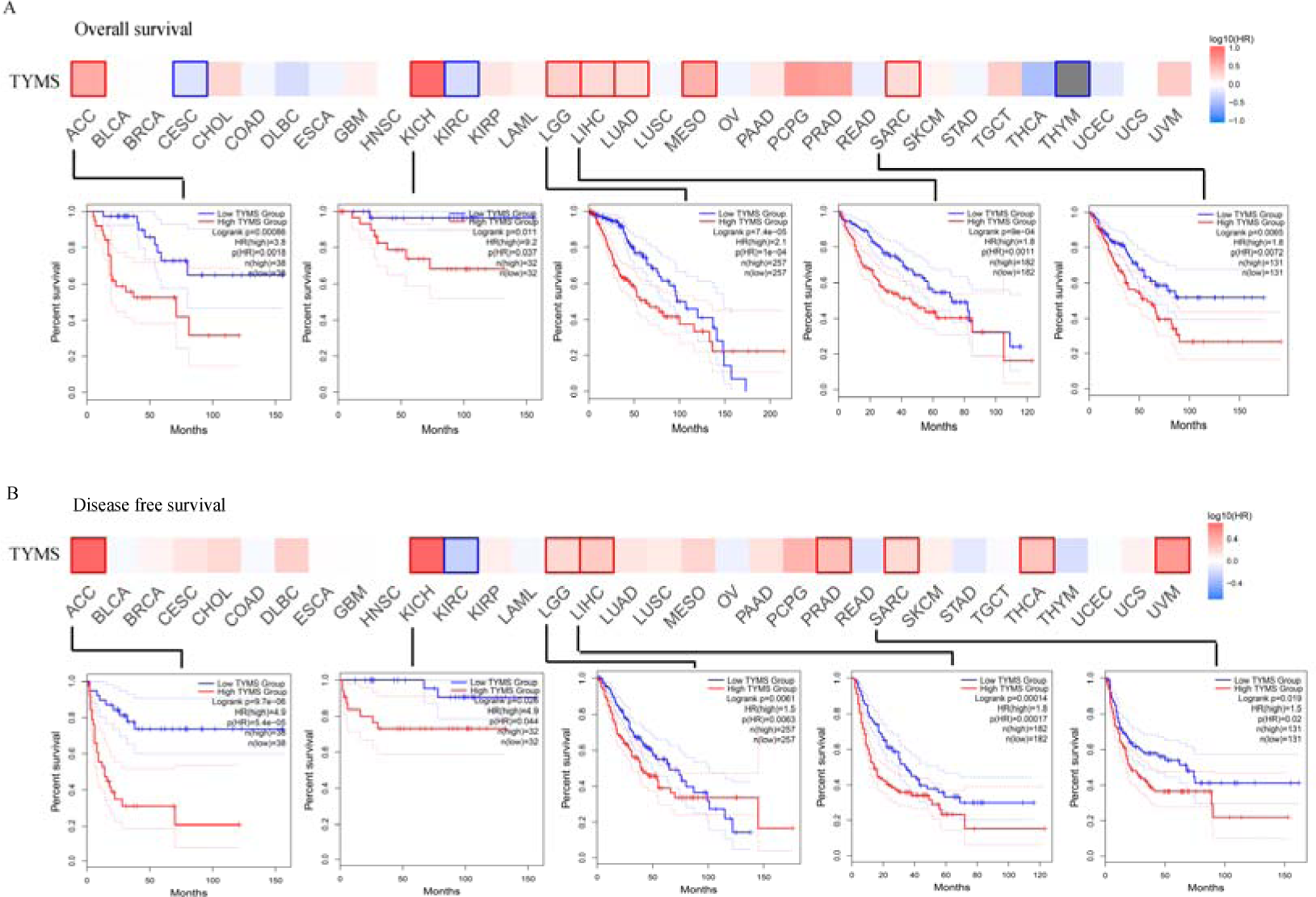
Correlation between TYMS expression and the survival outcome of tumors. OS (A) and DFS (B) was presented in heat bar. The significantly similar trend between OS was DFS was ACC, KICH, LGG, LIHC and SARC, which was displayed in survival map. (P < 0.05)

### Genetic alteration

The genetic alteration of TYMS has been observed in distinct tumors. The highest alteration frequency of TYMS (>4%) found in patients with bladder cancer was amplification (Figure 7A). The types, sites, and number of TYMS genetic alterations are depicted in Figure 7B. Overall survival in the TYMS alteration group was significantly shorter than in the wild type for both BLCA (P<0.01) and LUAD (P<0.05).

**Figure 7.**
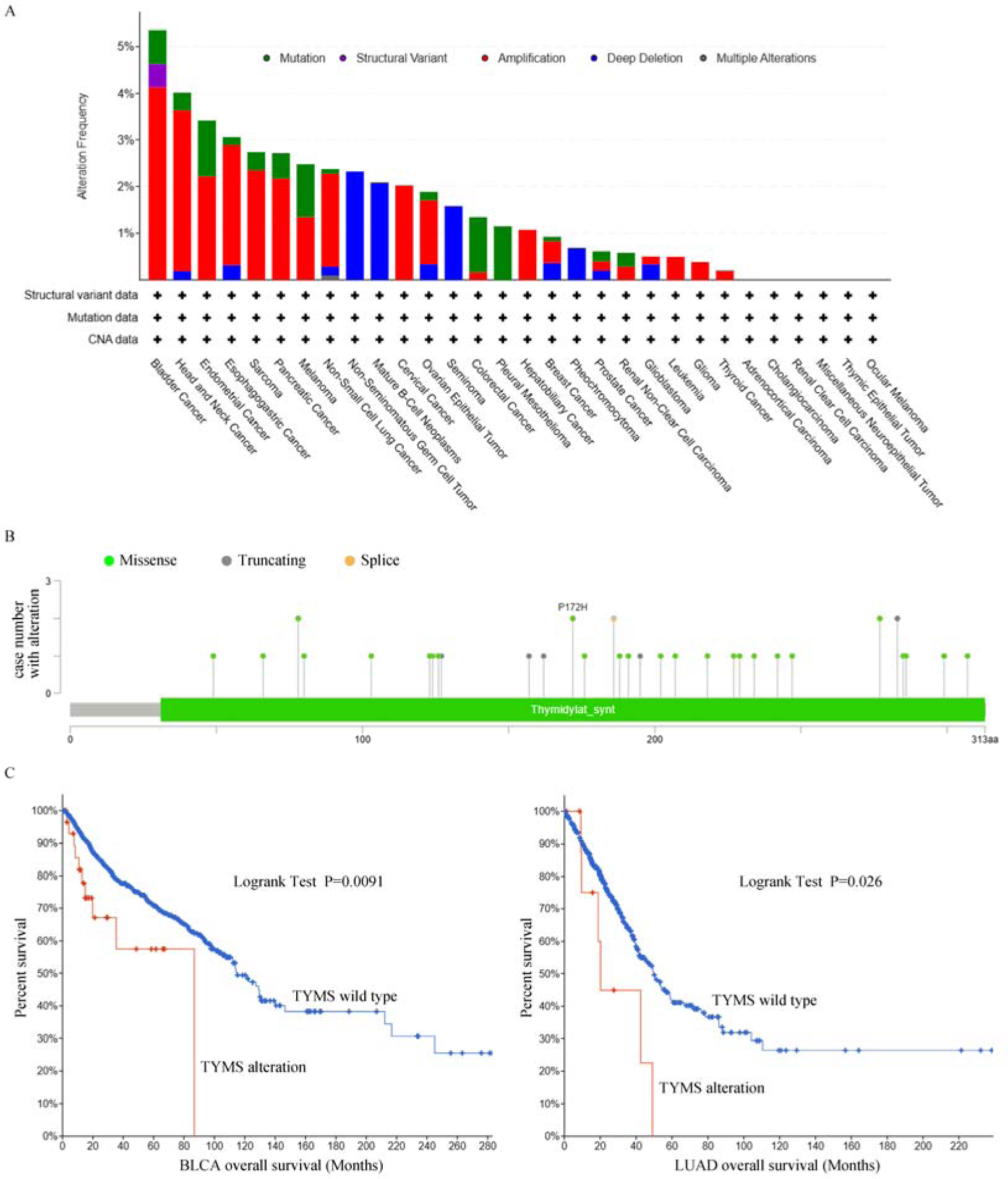
Mutation feature of TYMS. The alteration frequency with mutation type (A) and mutation site (B) is displayed. (C) There are significant overall survival differences between TYMS alteration and wild-type in BLCA and LUAD (P < 0.05).

### Phosphorylation analysis

Phosphorylation site analysis of TYMS in mammals was ahieved using the PhosphoNET database (Supplementary Figure 4). Unfortunately, limited studies explored phosphorylation differences between tumor and normal tissues, preventing a comprehensive summary of the implications of TYMS phosphorylation in tumors.

### Immune infiltration analysis

Tumor-infiltrating immune cells can enhance the development, progression, or metastasis of cancers[31]. A significant negative relationship was found between CD4+ T cells and TYMS expression in BLCA, DLBC, and MESO (Figure 8A). Additionally, a significant relationship between TYMS expression and cancer-associated fibroblasts was observed for BRCA, HNSC, KICH, STAD, TGCT, and THYM. Among them, BRCA, HNSC, STAD, and THYM exhibited a negative correlation (Figure 8B). However, similar trends were not observed in CD8+ and NK cell types (Supplementary Figure 5).

**Figure 8.**
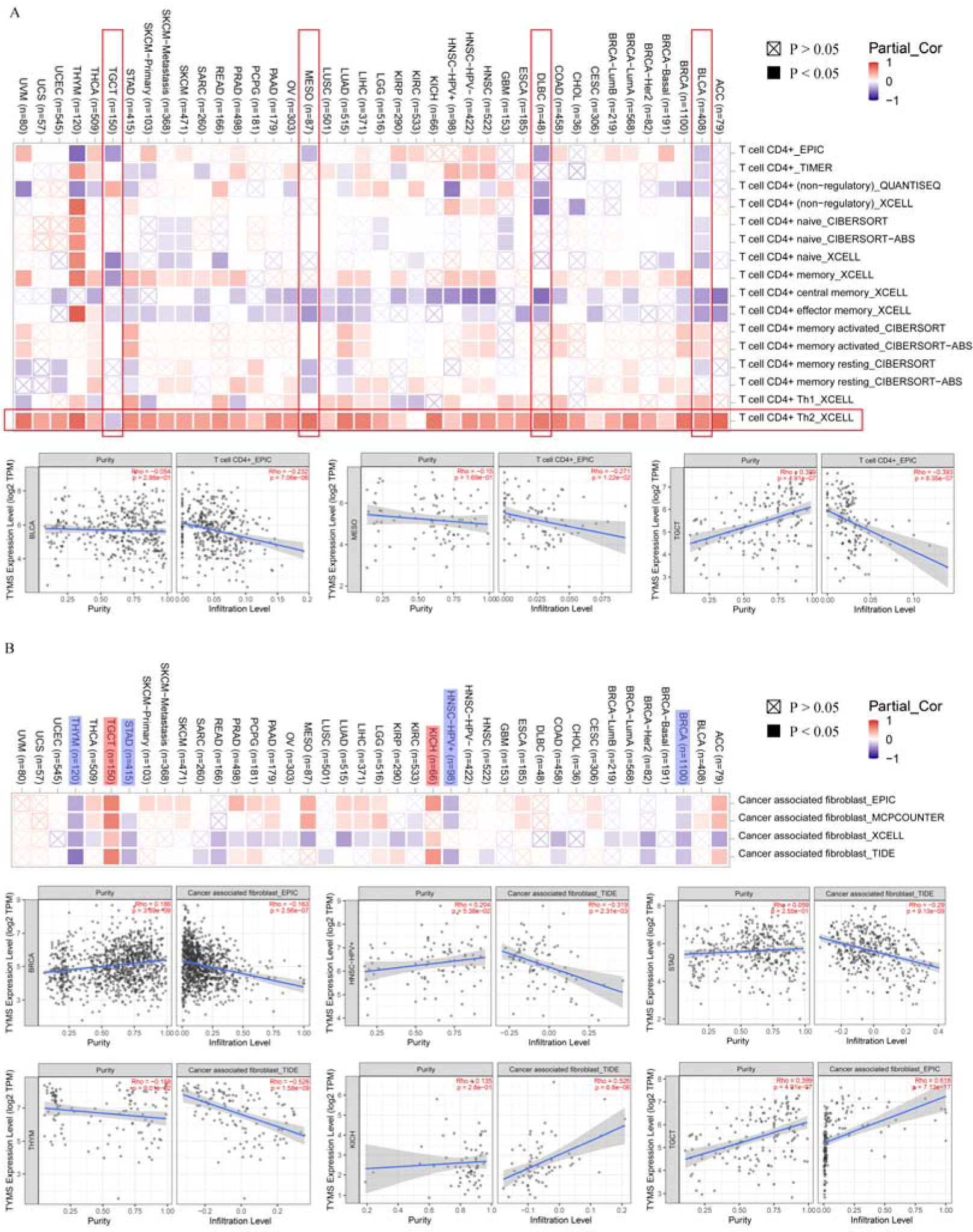
Correlation between TYMS and CD4+ immune cells (A) and cancer associated fibroblast (B).

### Enrichment analysis of TYMS-related genes

Fifty TYMS-binding proteins were identified through the STRING tool (Figure 9A). The top 100 TYMS-correlated targeting genes were summarized, with the top five being NDC80, EZH2, NUSAP1, WDR76, and MCM6. Notably, five genes overlapped in the two datasets (Figure 9B). The expression of these interacted genes (CDC45, MAD2L1, MCM5, NUSAP1, and PCAN) positively correlated with TYMS expression across almost all types of cancers (Figure 9C, D).

**Figure 9.**
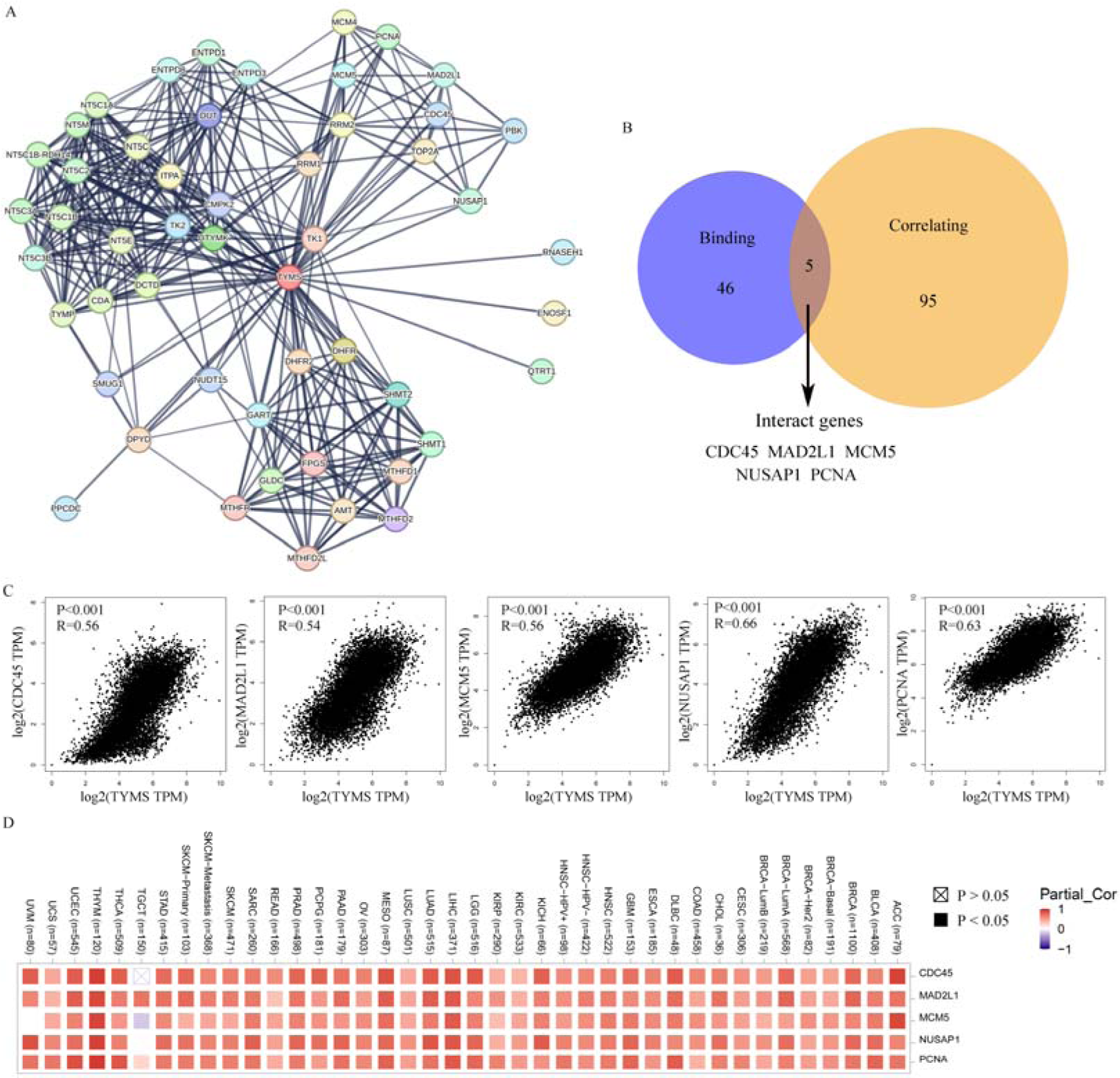
TYMS-related gene analysis. (A) Top 50 TYMS-binding proteins and their relationship. (B) The overlap part between the FASN-binding and correlated genes. (C) Expression analysis between TYMS and the five interacting genes in (B). (D) (C) displayed through heatmap across cancers.

Finally, GO analysis for these genes indicated their predominant location in the chromosomal region and execution of nucleotidase activity (Figure 10B, C). Furthermore, these genes were enriched in DNA replication, chromosome segregation, and nuclear division.

**Figure 10.**
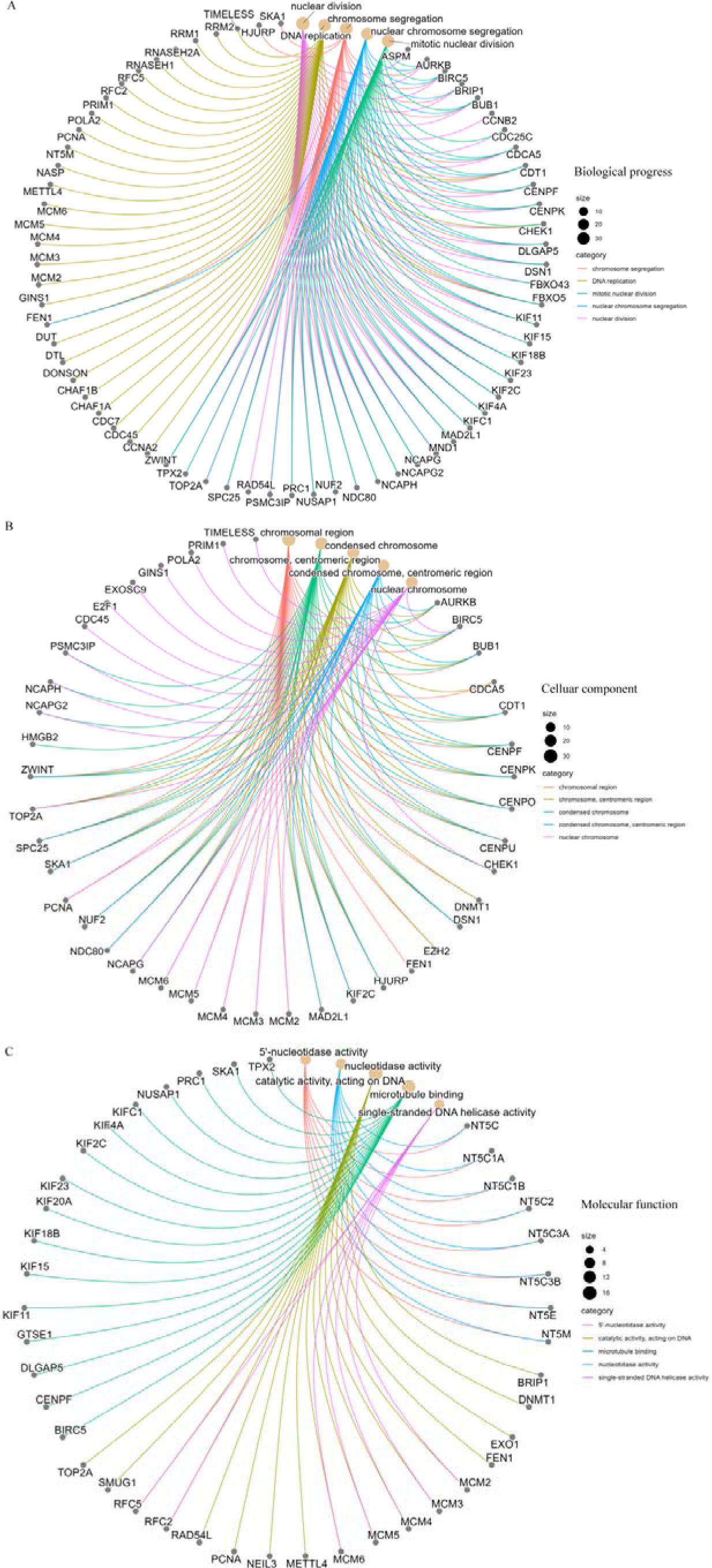
GO analysis of TYMS-related genes. Biological process (A), cellular component (B) and molecular function (C) was displayed.

## Discussion

TYMS, a folate-dependent essential enzyme, plays a pivotal role in generating intracellular de novo dTMP, critical for DNA synthesis and repair[32]. Its involvement extends beyond tumorigenesis, encompassing functions in coronary artery disease[33], virus replication and congenital disorder[34, 35]. This diversity underscores the significance of TYMS in various pathological contexts. The expanding literature linking TYMS to tumors prompted our comprehensive analysis to elucidate its oncogenic role in diverse cancer types.

The conservation of TYMS protein structure across species suggests shared physiological mechanisms. Notably, TYMS expression was detectable in most normal tissues and consistently elevated in tumor tissues compared to their normal counterparts. This widespread expression underscores the potential influence of TYMS in various cellular processes.

High TYMS expression consistently correlated with shorter overall survival and advanced tumor stage across multiple cancers. This association could be attributed to TYMS’s dual role: directly promoting tumor cell proliferation and inducing chemoresistance. Mechanistically, TYMS has been implicated in promoting cell proliferation and maintaining the de-differentiated state in breast cancer through distinct pathways[12, 36, 37]. Additionally, TYMS contributed to multiple drug resistance, impacting responses to 5-FU in COAD[6], temozolomide in glioma[38], pemetrexed in breast cancer/ MESO[39, 40], and platinum in NSCLC[41].

The interaction between TYMS and tumor-infiltrating immune cells highlights its role in the tumor microenvironment. A statistically significant difference was found between TYMS and CD4+ T-cells in BLCA, DLBC, MESO and TGCT (Figure 8A), which was similar with the previous studies. Dersh et al indicated that TYMS inhibitor increased MHC-1 presentation in DLBC, which revealed the role of TYMS in manipulating immunosurveillance in cancers[42]. Wang et al demonstrated that TYMS suppression impaired helper T cells (Th1 & Th17) differentiation and immune response[43]. Recently, a phase I clinical trial of thymidylate synthase poly-epitope-peptide vaccine against metastatic colorectal cancer induced immune-biological effects and showed antitumor activity[44]. Also, a positive correlation between TYMS and cancer-associated fibroblast in KICH and TGCT which need further investigations. Above all, these results provided evidence that TYMS have a complicated correlation with immune system and deserved further studies in tumor immunology and microenvironment.

Enrichment analysis combining TYMS-binding components and expression-related genes revealed the central involvement of “nucleotidase activity” and “DNA replication” in tumor cell proliferation. These processes are pivotal in the progression of tumors, emphasizing the need for further experimental validation to elucidate the specific oncogenic role of TYMS. Future studies should delve into the molecular mechanisms underlying TYMS-mediated tumor progression and its potential as a therapeutic target.

## Conclusion

Based on a comprehensive analysis across various tumors, we found a factual association between TYMS expression and clinical outcome, protein phosphorylation and immune cell infiltration, as well as relating genes and functions, which could help to better understand the oncogenic role of TYMS.

## Author contribution

YG performed the most data analysis and assisted with the writing of the manuscript; LX performed the rest data analysis; YG, Yang W and Yan W designed the study and assisted with the writing of the manuscript; YG and Yan W performed the literature search and collected the data; Yan W supervised the study; All authors have read and approved the manuscript.

## Data Availability

All data produced in the present study are available upon reasonable request to the authors

## Abbreviations

ACC: Adrenocortical carcinoma
BLCA: Bladder Urothelial Carcinoma
BRCA: Breast invasive carcinoma Cervical squamous cell carcinoma and endocervical
CESC: adenocarcinoma
CHOL: Cholangiocarcinoma
COAD: Colon adenocarcinoma
DFS: Disease free survival
DLBC: Diffuse Large B-cell Lymphoma
dTMP: deoxythymidine monophosphate
ESCA: Esophageal carcinoma
GBM: Glioblastoma
GEPIA2: Gene Expression Profiling Interactive Analysis, version 2
GTEx: Genotype-Tissue Expression
HNSC: Head and Neck squamous cell carcinoma
HPA: The Human Protein Atlas
IHC: Immunohistochemistry
KICH: Kidney Chromophobe
KIRC: Kidney renal clear cell carcinoma
KIRP: Kidney renal papillary cell carcinoma
LAML: Acute Myeloid Leukemia
LGG: Low grade glioma
LIHC: Liver hepatocellular carcinoma
LUAD: Lung adenocarcinoma
LUSC: Lung squamous cell carcinoma
MESO: Mesothelioma
MHC: major histocompatibility complex
NCBI: National Center for Biotechnology Information
NK: Natural killer
NSCLC: non-small cell lung carcinoma
OS: Overall survival
OV: Ovarian serous cystadenocarcinoma
PAAD: Pancreatic adenocarcinoma
PCPG: Pheochromocytoma and Paraganglioma
PRAD: Prostate adenocarcinoma
READ: Prostate adenocarcinoma
SARC: Sarcoma
SKCM: Skin Cutaneous Melanoma
STAD: Stomach adenocarcinoma
TCGA: The cancer genome atlas
TGCT: Testicular Germ Cell Tumors
THCA: Testicular Germ Cell Tumors
THYM: Thymoma
TIMER2: tumor immune estimation resource, version 2
TPM: Transcripts per million
TYMS: thymidylate synthase
UCEC: Uterine Corpus Endometrial Carcinoma
UCS: Uterine Carcinosarcoma
UVM: Uveal Melanoma

## Acknowledgements

This work was supported by the Beijing Municipal Natural Science Foundation (No. 7244344), Beijing Chao-yang Hospital Golden Seed foundation (No. CYJZ202201) and the inner-hospital foundation of Beijing Chao-yang Hospital. Besides, we would like to thank Xulei Huo for his technical supports.

---

**Supplementary Figure 1.** Structural characteristics of TYMS. (A) Gene location of TYMS. (B) Conserved domain of TYMS. (C) Three-dimension structure of TYMS.

**Supplementary Figure 2.** TYMS expression showed no difference between KICH, PRAD, TGCT and THCA and their comparable normal tissues.

**Supplementary Figure 3.** TYMS expression across pathological stages showed no difference in these tumors.

**Supplementary Figure 4.** Phosphoprotein site of TYMS showed in the axis Supplementary Figure 5. Correlation between TYMS and CD8+ immune cells (A) and NK cells (B).

**Supplementary File 1.** The reference list related to TYMS phosphoprotein site in mammals.

**Supplementary Table 1.** Top 100 TYMS-correlated genes

